# Explaining ethnic disparities in COVID-19 mortality: population-based, prospective cohort study

**DOI:** 10.1101/2021.02.07.21251079

**Authors:** G. David Batty, Bamba Gaye, Catharine R Gale, Mark Hamer, Camille Lassale

## Abstract

Ethnic disparities in COVID-19 hospitalizations and mortality have been reported but there is scant understanding of how these inequalities are embodied. The UK Biobank prospective cohort study comprises around half a million people who were aged 40-69 years at study induction between 2006 and 2010 when information on ethnic background and potential explanatory factors was captured. Study members were linked to a national mortality registry. In an analytical sample of 448,664 individuals (248,820 women), 354 deaths were ascribed to COVID-19 between 5^th^ March and the end of follow-up on 17^th^ September 2020. In age- and sex-adjusted analyses, relative to White participants, Black study members experienced around seven times the risk of COVID-19 mortality (odds ratio; 95% confidence interval: 7.25; 4.65, 11.33), while there was a doubling in the Asian group (1.98; 1.02, 3.84). Controlling for baseline comorbidities, socioeconomic circumstances, and lifestyle factors explained 53% of the differential in risk for Asian people (1.37; 0.68, 2.77) and 27% in Black study members (4.28; 2.67, 6.86). The residual risk in ethnic minority groups for COVID-19 deaths may be ascribed to unknown genetic factors or unmeasured phenotypes, most obviously racial discrimination.

## Introduction

Although the 2009 swine influenza (H1N1) pandemic did not have the acute and far-reaching societal and economic impact of coronavirus disease 2019 (COVID-19), severe cases were nonetheless characterised by ethnic disparities.^1-3^ In the present pandemic, there is also emerging evidence of such inequalities whereby, relative to white individuals, people of Afro-Caribbean (Black), Latinx, and, to a lesser extent, Asian origin, experience the greatest burden of severe acute respiratory syndrome coronavirus 2 (SARS-CoV-2) – the infection that causes COVID-19 – and hospitalization for, and mortality from, this disease.^4-6 7^

Understanding how these ethnic variations in COVID-19 are embodied is central to the process of disease prevention. Individuals from different ethnic backgrounds vary in behaviours, body composition, comorbidities, immune profiles, and socioeconomic circumstances,^8^ yet with studies in this field typically generated from electronic health records^9-12^ these potential explanatory factors are rarely measured. For instance, a recent systematic review on ethnicity and COVID-19 largely located such electronic records-based studies,^13^ and revealed that, while investigators generally took into account socioeconomic status and somatic morbidities, statistical control for other important phenotypes such as mental health,^14^ lifestyle factors (e.g., body mass index, alcohol intake),^15^ and physiological indices (e.g., systemic inflammation)^16,17^ was lacking.

Using data from UK Biobank, a field-based prospective cohort study, we have shown that people of Asian and particularly Afro-Caribbean (Black) heritage experienced a markedly elevated risk of a severe COVID-19 diagnosis, and up to half of these differentials was explained by socioeconomic and lifestyle indices.^18^ In that study, hospitalisation for COVID-19 was the outcome of interest. As the pandemic has unfolded, sufficient deaths from the disease have accumulated to allow us to test these original results with new data.

## Methods

UK Biobank is a prospective cohort study, the sampling and procedures of which have been well described.^19^ Baseline data collection took place between 2006 and 2010 across twenty two research assessment centres in the UK giving rise to a sample of 502,655 people aged 40 to 69 years (response rate 5.5%). Ethical approval was granted by the North-West Multi-centre Research Ethics Committee, and the research was carried out in accordance with the Declaration of Helsinki of the World Medical Association; participants gave written consent.

Ethnicity was self-classified as White (British, Irish, any other white background); Asian or Asian British (Indian, Pakistani, Bangladeshi, any other Asian background); Black or Black British (Caribbean, African, any other Black background); Mixed; Chinese; or ‘other’.^18^ With a low number of COVID-19 deaths occurring in the latter three categories, these were collapsed into a single ‘other’ group. Individual socioeconomic status was captured using educational qualifications (university degree, other qualifications, no qualifications), and the number of people in the household of the study member (living alone, 2 people, 3 people, 4 people or more).^20^ A third indicator of individual socioeconomic status – occupational classification – was available in a subgroup of participants (N=322,353) and based on current job from which we derived two categories: non-manual (managerial positions, technical, administrative) and manual (sales and customer service, process, plant & machine operatives). The Townsend index of neighbourhood deprivation is based on national census data, with each participant assigned a score corresponding to the postcode of home address; scores were categorised into quintiles such that higher values denoted greater disadvantage (≤ −3.960; −3.959 to −2.828; −2.827 to −1.414; −1.413 to +1.140; ≥ 1.141). Levels of cigarette smoking (never, former, current) and alcohol consumption (never, special occasions, 1-3/month, 1-2 week, 3-4/week, daily) were assessed using standard enquiries. Height, weight, and circumferences of waist and hip were measured using standard protocols. Vascular or heart problems, diabetes, and chronic bronchitis, were based on self-reported physician diagnosis and presence of hypertension was defined as systolic/diastolic blood pressure ≥ 140/90 mmHg and/or self-reported use of antihypertensive medication. Study members were also asked whether they had ever been under the care of a psychiatrist for any mental health problem.^14^ Available for a subgroup (N=358,820), white blood cell count, glycated haemoglobin, and high-density lipoprotein cholesterol concentrations were based on assays of non-fasting venous blood. Participants were linked to long-standing national mortality records from which death from COVID-19, our outcome of interest, was denoted by the emergency International Classification of Disease (version 10) code U07.1 (COVID-19, virus identified).

### Statistical analyses

To summarise the association of mortality with ethnicity we used logistic regression to compute odds ratios with accompanying 95% confidence intervals. With COVID-19 deaths occurring over a short period and being rare in the present study, odds ratios closely resemble hazard ratios as computed using Cox regression analyses.

## Results

In an analytical sample of 448,664 individuals (248,820 women), 354 deaths were ascribed to COVID-19 between 5^th^ March and the end of follow-up on 17^th^ September 2020 (320 in White participants; 21 in Blacks; 9 in Asians; and 4 in those from other ethnic groups). In table 1, we show age- and sex-adjusted odds ratios for both ethnicity and covariates in relation to the risk of death from COVID-19. Unfavourable levels of all fourteen covariates were related to a higher risk of death from COVID-19 in minimally-adjusted analyses; only the point estimate for chronic bronchitis, while elevated, did not achieve statistical significance at conventional levels. Thus, there was a raised risk of COVID-19 death in people from disadvantaged socioeconomic background and those with extant illness at baseline. While people with less healthy lifestyle choices typically experienced higher risk, the daily consumption of alcohol seemed to confer protection. Relative to white participants, Black study members experienced around seven times the risk of COVID-19 mortality (age- and sex-adjusted odds ratio; 95% confidence interval: 7.25; 4.65, 11.33), while there was around a doubling in the Asian group (1.98; 1.02, 3.84). There was evidence of a lack of precision in some of these analyses as evidenced by the breadth of the confidence intervals.

**Table 1.**
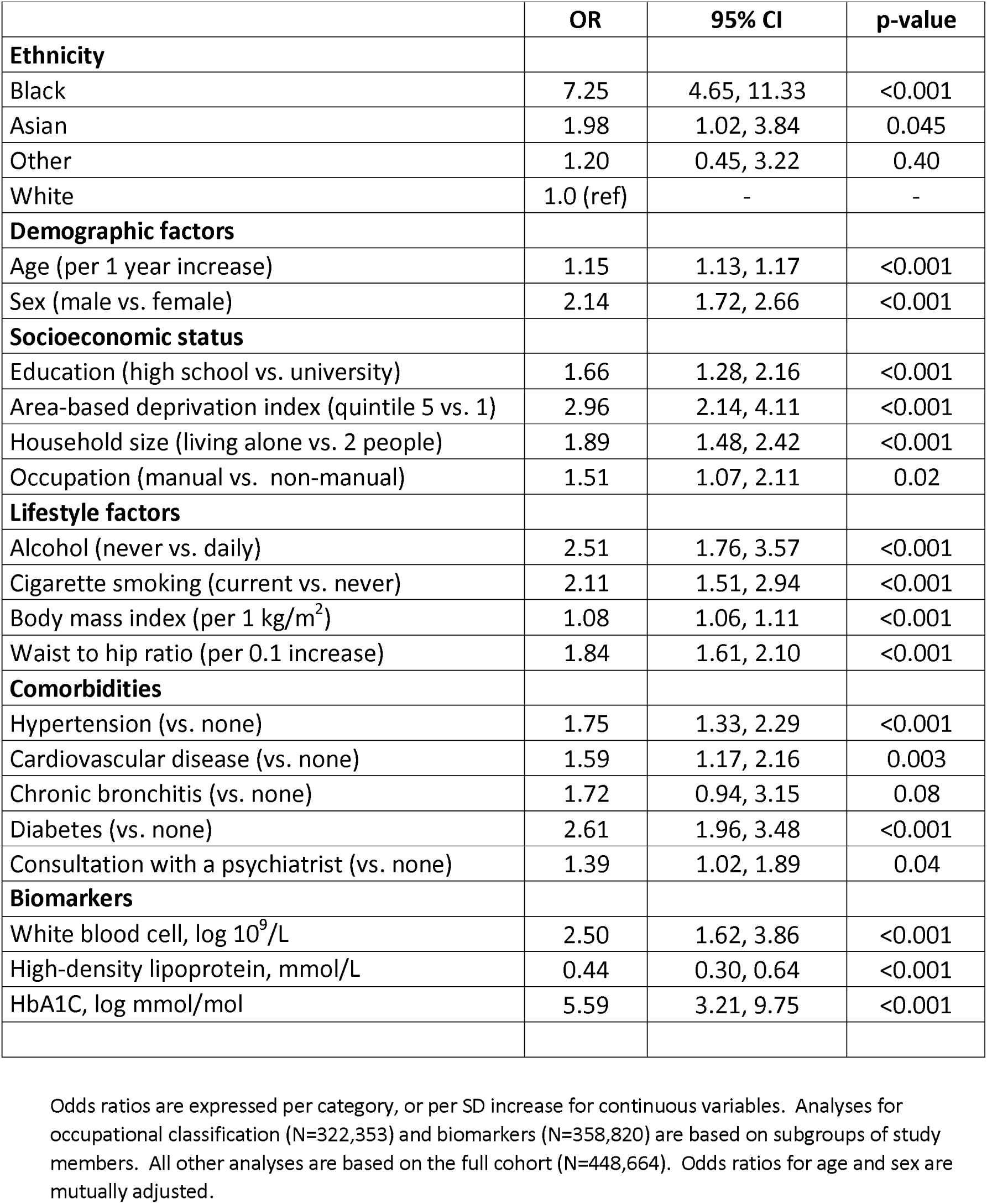
Age- and sex-adjusted odds ratios for the association of ethnicity and baseline covariates (2006-10) with COVID-19 mortality (2020)

|  | **OR** | **95% CI** | **p-value** |
| --- | --- | --- | --- |
| **Ethnicity** |  |  |  |
| Black | 7.25 | 4.65, 11.33 | <0.001 |
| Asian | 1.98 | 1.02, 3.84 | 0.045 |
| Other | 1.20 | 0.45, 3.22 | 0.40 |
| White | 1.0 (ref) | - | - |
| **Demographic factors** |  |  |  |
| Age (per 1 year increase) | 1.15 | 1.13, 1.17 | <0.001 |
| Sex (male vs. female) | 2.14 | 1.72, 2.66 | <0.001 |
| **Socioeconomic status** |  |  |  |
| Education (high school vs. university) | 1.66 | 1.28, 2.16 | <0.001 |
| Area-based deprivation index (quintile 5 vs. 1) | 2.96 | 2.14, 4.11 | <0.001 |
| Household size (living alone vs. 2 people) | 1.89 | 1.48, 2.42 | <0.001 |
| Occupation (manual vs. non-manual) | 1.51 | 1.07, 2.11 | 0.02 |
| **Lifestyle factors** |  |  |  |
| Alcohol (never vs. daily) | 2.51 | 1.76, 3.57 | <0.001 |
| Cigarette smoking (current vs. never) | 2.11 | 1.51, 2.94 | <0.001 |
| Body mass index (per 1 kg/m<sup>2</sup>) | 1.08 | 1.06, 1.11 | <0.001 |
| Waist to hip ratio (per 0.1 increase) | 1.84 | 1.61, 2.10 | <0.001 |
| **Comorbidities** |  |  |  |
| Hypertension (vs. none) | 1.75 | 1.33, 2.29 | <0.001 |
| Cardiovascular disease (vs. none) | 1.59 | 1.17, 2.16 | 0.003 |
| Chronic bronchitis (vs. none) | 1.72 | 0.94, 3.15 | 0.08 |
| Diabetes (vs. none) | 2.61 | 1.96, 3.48 | <0.001 |
| Consultation with a psychiatrist (vs. none) | 1.39 | 1.02, 1.89 | 0.04 |
| **Biomarkers** |  |  |  |
| White blood cell, log 10<sup>9</sup>/L | 2.50 | 1.62, 3.86 | <0.001 |
| High-density lipoprotein, mmol/L | 0.44 | 0.30, 0.64 | <0.001 |
| HbA1C, log mmol/mol | 5.59 | 3.21, 9.75 | <0.001 |

Odds ratios are expressed per category, or per SD increase for continuous variables. Analyses for occupational classification (N=322,353) and biomarkers (N=358,820) are based on subgroups of study members. All other analyses are based on the full cohort (N=448,664). Odds ratios for age and sex are mutually adjusted.

We explored the impact of individual covariates by making separate (non-accumulative) adjustment for socioeconomic indices, lifestyle factors, and comorbidities (figure 1 and aTable 1, appendix). In Black participants, relative to the regression coefficients in the age- and sex-adjusted analyses, we found that socioeconomic factors offered the most explanatory power (4.61; 2.91, 7.32; 23% attenuation), whereas in people of Asian backgrounds, it was lifestyle factors (1.56; 0.79, 3.09; 34% attenuation) and comorbidities (1.61; 0.82, 3.15; 30% attenuation). Collectively, these covariates explained more of the disparities for Asian (1.37; 0.68, 2.77; 53% attenuation) than for black individuals (4.29; 2.67, 6.88; 27% attenuation).

**Figure 1.**
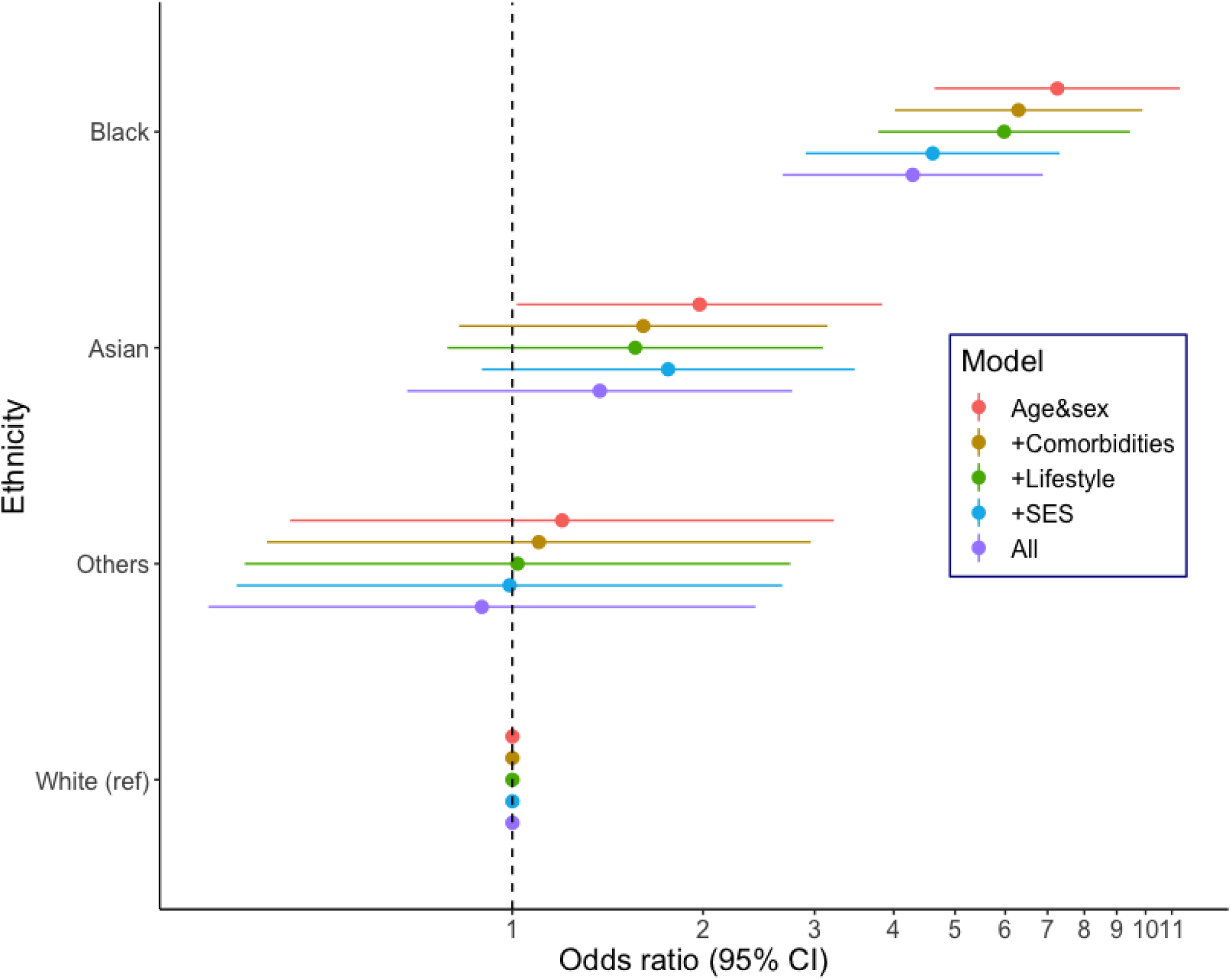
Odds ratios for the association between ethnicity and COVID-19 mortality. Covariates included in each model correspond to those described in table 1. For the black group, attenuation of regression coefficients was: 22.9% after controlling for socioeconomic status; 9.8% for lifestyle; 7.1% for comorbidity; and 26.5% for all covariates combined. For the Asian group: 16.8% after controlling for socioeconomic status; 34.3% for lifestyle; 30.0% for comorbidities; and 53.3% for all covariates combined. For the ‘other’ ethnic group: 105.6% after controlling for socioeconomic status; 89.6% for lifestyle; 46.7% for comorbidities; and 161.4% for all covariates combined.

That we were only able to partially explain ethnic inequalities in the present analyses raises the issue of their being a role for other phenotypic risk indices. Data on biological risk factors were captured in a subgroup of study participants. Biological indices including high density lipoprotein cholesterol, glycated haemoglobin, and white blood cell count, associated with COVID-19 deaths in the present dataset, were available for 358,820 people in whom there were 290 COVID-19 deaths (266 in Whites; 13 in Blacks; 7 in Asians; and 4 in people from other ethnic groups). Adding these variables to the basic statistical model yielded marked attenuation (1.47; 0.69, 3.14; 43% attenuation) for Asian study members but not for Black individuals (6.56; 3.69, 11.69; 1% attenuation) (aTable 2, appendix).

Lastly, it is plausible that people with ethnic minority ancestry are more likely to be in service industry employment which requires them to have a person- or patient-facing role so elevating their risk of infection. In analyses of the subgroup with data on job title (N=322,353) there was a total of 177 deaths (156 in Whites; 15 in Blacks; 4 in Asians, and 2 in other). The raised risk in Black individuals was in fact elevated marginally more (5.55; 3.17, 9.73) when occupation was added to the original adjustment for socioeconomic status in the main analyses (4.27; 2.40, 7.60); again, statistical precision was modest owing to the small numbers of COVID-19 fatalities (aTable3, appendix).

## Discussion

Our main finding was that the elevated risk of COVID-19 mortality in people from ethnic minority backgrounds was partially explained by socioeconomic indices, lifestyle factors, and comorbidities. That is, relative to the white group, these factors collectively explained around one quarter of the raised risk in people of Black origin and around half in those of Asian ancestry. That we were able to replicate known associations with COVID-19 mortality for socioeconomic circumstances, comorbidities, age, and sex from US,^21^ UK,^22^ Italy,^23^ China,^24,25^ and Brazil^26^ in the present dataset gives us some confidence in the more novel results presented here for ethnicity.

The residual risk of COVID-19 in black study members suggests that genetic and/or unmeasured environmental factors also have a role.^8^ Such genetic factors are currently unknown, and while the present dataset is reasonably well-characterised for environmental factors, it lacks data on racial discrimination which appears to have an influence on selected health outcomes, most consistently mental health.^27^ Whether racial discrimination is involved in the occurrence of COVID-19 is plausible given links to other respiratory conditions – elevated rates of adult-onset asthma are apparent in people who report experiencing higher levels of everyday racism^28^ – and has been vigorously advanced as having a role in the current pandemic.^29,30^ To the best of our knowledge, this link has yet to be tested empirically.

### Comparison with existing studies

Although less well examined owing to lower societal burden, the 2009 H1N1 pandemic revealed similar ethnic differentials to those reported herein ^1,2^ The Spanish influenza of 1918 was perhaps an exception: rates of hospitalisations and death were in fact seemingly lower in people of Black ethnic origin relative to Whites in the US^31^ – the only year in the 20th century when being of Black origin appeared to confer some protection against death from influenza. In the current pandemic, the present findings of ethnic disparities are supported by observations made on populations from the US^4^ and the UK.^5,6^ While, as discussed, in-depth examination of the causes of these inequalities is rare owing to an absence of higher resolution data in most studies, effects seem to survive adjustment for extant morbidity and, when available, markers of poverty.^9-12^ Partial attention by comorbidity, which for the first time to our knowledge featured mental illness,^14^ was also seen herein, with additional explanatory power offered by lifestyle factors and socio-economic circumstances which confirms our earlier work based on for hospitalisations for the disease.^18^ Unlike the present analyses featuring death as the outcome of interest, in that study^18^ we used a record of a positive in-patient test for COVID-19 as our outcome of interest. While this was assumed to be an indicator of disease severity – only serious cases are hospitalised in the UK which operates under a single, national health service – it is nonetheless plausible that some of the cases were patients being treated for unrelated conditions who were asymptomatically positive for COVID-19 after routine hospital-wide testing. Our results here for death from the disease for ethnicity therefore corroborates these earlier findings.^18^

### Study strengths and weaknesses

The strengths of the study include the well-characterised nature of the study members and the full coverage of the population for cause of death from COVID-19. The study is of course not without its weaknesses. Although the present cohort is large, there were too few deaths in selected ethnic groups – people from Chinese or mixed backgrounds, for instance – to facilitate analyses. With the present sample not being representative of the general UK population, death rates from leading causes and the prevalence of reported risk factors are known to be underestimates of those apparent in less select groups;^32^ the same is likely to be the case for COVID-19 cases. This notwithstanding, there is evidence that risk factor associations, including those presented herein, are externally valid.^32^ Lastly, while ethnicity itself is stable over-time – UK data reveal that only 4% of census participants chose a different ethnic group a decade after their first declaration^33^ – other baseline data are more likely to be time-varying in the period between study induction in UK Biobank and the present pandemic. This is a perennial issue in cohort studies and one we were able to investigate using data from a resurvey that took place around 8 years after baseline examination in a sub-sample. Analyses revealed moderate to high stability for covariates central to the present analyses, including education (r=0.86, p<0.001, N=30,350), cigarette smoking (r=0.60, p<0.001, N=31037), and body mass index (r=0.90, p<0.001, N=34,662).

In conclusion, in this well-characterised prospective cohort study, we were only able to partially explain how ethnic disparities in COVID-19 were embodied. The residual risk in COVID-19 deaths may be ascribed to systemic racism and other unmeasured phenotypic or genetic factors.

## Data Availability

Data from UK Biobank (http://www.ukbiobank.ac.uk/) are available to bona fide researchers upon application.

